# Within-Group Racial and Ethnic Differences in County-Level Socio-Behavioral Risk Across Cancer Mortality Tertiles in the United States

**DOI:** 10.64898/2026.02.24.26347030

**Authors:** Valerie C. Valerio, Talita Honorato-Rzeszewicz, Peter Smittenaar, Sema K. Sgaier, Camille Jimenez

## Abstract

**Importance:** Persistent racial and ethnic disparities in breast and prostate cancer mortality are well documented. Most prior studies emphasize between-group differences and rely on population averages or single composite measures of social disadvantage, which can obscure high-need communities within groups. How socio-behavioral determinants of health vary within groups across local gradients of cancer mortality remains incompletely characterized. A framework that combines race- and cancer-specific mortality with local, domain-level socio-behavioral profiles may help identify where burden is greatest and which specific barriers warrant prioritization.

**Objective:** To determine how socio-behavioral risk relates to breast and prostate cancer mortality within racial and ethnic groups and to characterize domain-specific behavioral profiles across low-, moderate- and high-mortality counties to inform targeted, equity-oriented cancer control strategies.

**Design:** Cross-sectional study of U.S. counties.

**Setting:** United States, county-level analysis.

**Participants:** 3,141 U.S. counties, stratified within Non-Hispanic White, Non-Hispanic Black, and Hispanic populations.

**Exposures:** County-level socio-behavioral determinants of health measured using a composite index comprising seven domains: community solidarity; education, health literacy, and digital connectivity; quality of care; housing and environmental risk; economic livelihoods; lifestyle behaviors; and touchpoints with care.

**Main outcomes and measures:** Race/ethnicity-specific, age-adjusted breast and prostate cancer mortality rates (2018-2022) and county-level socio-behavioral risk scores. Counties were grouped into mortality tertiles within each race/ethnicity-by-cancer-stratum.

**Results:** Across groups, higher socio-behavioral risk was associated with higher breast and prostate cancer mortality. For breast cancer, socio-behavioral risk increased monotonically across mortality tertiles for all groups, with the largest within-group increases among Hispanic and Non-Hispanic Black women. For prostate cancer, risk generally increased across mortality tertiles for all groups. Although Hispanic populations had lower population-average mortality, high-mortality Hispanic counties exhibited pronounced risk in lifestyle behaviors, economic livelihoods, and touchpoints with care. Domain patterns associated with high mortality varied by race, ethnicity, and cancer type, with touchpoints with care and economic livelihoods consistently prominent.

**Conclusions and relevance:** Within-group heterogeneity in socio-behavioral risk is substantial across U.S. counties. Linking population-specific, domain-level socio-behavioral profiles to cancer mortality may support more precise and equity-oriented cancer control strategies than reliance on group averages or composite indices.

**Key points:** *Question:* Within racial and ethnic groups, how do socio-behavioral determinants of health vary across US counties with low, moderate, and high breast and prostate cancer mortality?

*Findings:* In this cross-sectional study, higher county-level socio-behavioral risk was associated with higher breast and prostate cancer mortality across racial and ethnic groups. Race/ethnicity-specific, domain-level profiles revealed within-group heterogeneity, including persistently elevated risk among Non-Hispanic Black populations and pronounced domain-specific gaps in high-mortality Hispanic counties.

*Meaning:* Linking population-specific socio-behavioral profiles to local cancer mortality can guide more precise and equity-oriented prioritization of intervention domains and geographies than reliance on group averages or composite indices.

## Introduction

Breast and prostate cancer are among the most frequently diagnosed cancers in the United States and are leading causes of cancer-related mortality (1–3). Breast and prostate cancer are common across the life course (1,2). Despite advances in screening and treatment, substantial disparities in cancer outcomes persist across race, ethnicity, and geography. Black and Hispanic women are less likely to be diagnosed at an early stage and experience lower breast cancer survival than White women (4), while prostate cancer mortality among Black men is approximately twice that among White men (3,5). These patterns are consistently observed in SEER data (4).

Socio-behavioral determinants of health (SBDOH), an extension of the social determinants of health framework that integrates structural social conditions with population-level health-related behaviors and care engagement, are associated with cancer incidence, stage at diagnosis, treatment access, and survival (6–9). Prior studies have demonstrated that geographic variation in these determinants corresponds closely with county-level patterns of breast and prostate cancer mortality in the United States (7,9). Because minority populations are more likely to be exposed to adverse socio-behavioral and structural conditions, unequal distributions of SBDOH contribute to observed disparities in cancer outcomes (6,8).

However, socio-behavioral risk is heterogeneous and may arise from different combinations of economic, structural, cultural, and environmental factors. Individuals or communities with similar overall levels of vulnerability may face distinct barriers to prevention, early detection, and treatment. While previous studies have documented racial, ethnic, and geographic disparities in cancer outcomes and their associations with SBDOH, few have examined how socio-behavioral needs vary within racial and ethnic groups across gradients of cancer mortality risk.

In this study, we assess and compare the socio-behavioral needs of Non-Hispanic White, Non-Hispanic Black, and Hispanic populations residing in U.S. counties with low, moderate, and high breast and prostate cancer mortality risk using standard race and ethnicity definitions (10,11). By characterizing differentiated socio-behavioral profiles within racial and ethnic groups across mortality-risk strata, this study aims to inform more precise, population-tailored, and equity-oriented cancer prevention, screening, and survivorship strategies.

## Methods

### Definitions

We define socio-behavioral determinants of health as structural conditions combined with population-level health-related behaviors and care engagement. We operationalize socio-behavioral risk as higher values on the socio-behavioral determinants of health index, with higher scores indicating a greater prevalence of adverse socio-behavioral determinants. We use *need* to denote areas where socio-behavioral risk is highest and greater investment may be warranted; this does not imply individual-level clinical need.

### Study design and population

This cross-sectional study analyzed county-level breast and prostate cancer mortality, socio-behavioral determinants of health (SBDOH), and race/ethnicity-specific population estimates for Non-Hispanic White, Non-Hispanic Black, and Hispanic populations. Race and ethnicity were classified according to the Census Bureau definitions (see eMethods for details). The unit of analysis was the county.

### Cancer mortality data

Age-adjusted breast and prostate cancer mortality rates stratified by race and ethnicity for the period 2018-2022 were obtained from the National Cancer Institute (4). Mortality rates were expressed per 100,000 population and age-adjusted to the 2000 U.S. standard population. To protect patient confidentiality, mortality rates were suppressed for counties with fewer than 10 deaths per year. Counties with suppressed mortality rates were excluded within each race/ethnicity-by-cancer stratum.

### Socio-behavioral determinants of health data

County-level SBDOH indicators and race/ethnicity-specific population estimates were derived from multiple sources, including the American Community Survey (ACS) and other national data sources. The most recent year of data available for each indicator was used.

We included 59 indicators across 7 domains: community solidarity (9 indicators), education, health literacy, and digital connectivity (8 indicators), quality of care (7 indicators), housing and environmental risks (11 indicators), economic livelihoods (9 indicators), lifestyle behaviors (6 indicators), and touchpoints with care (9 indicators). A list of all indicators is provided in eTable 1.

### Construction of race/ethnicity-specific SBDOH scores

We constructed race/ethnicity-specific census tract-level SBDOH measures from tract-level indicators. Tract-level values were aggregated to the county level using population-weighted averaging by race and ethnicity (see eMethods).

For each county and racial/ethnic group, relative SBDOH risk scores were constructed using percentile ranking (12). Each county received one overall SBDOH index score, and 7 domain-specific thematic scores, with all scores scaled from 0 (lowest risk) to 100 (highest risk).

### Classification of counties by cancer mortality risk

For each racial/ethnic group and cancer type, counties were classified into mortality tertiles (low, moderate, and high) based on race/ethnicity-specific age-adjusted mortality rates per 100,000 population. Tertiles were generated separately for 1) Female adult populations for breast cancer, and 2) Male adult populations for prostate cancer, within each racial and ethnic group. Counties with missing or suppressed mortality data were excluded from their respective analysis. The total represented population within each tertile was quantified for each racial/ethnic group and cancer type. Socio-behavioral data were available for all counties included in the mortality analyses.

### Statistical analysis

Using county-level socio-behavioral determinants of health scores and sex-specific adult population counts from the American Community Survey, we calculated population-weighted mean socio-behavioral risk scores across cancer mortality tertiles for each racial and ethnic group and cancer type.

Because distributions were nonnormal, we used non-parametric methods. Differences in socio-behavioral risk scores across low-, moderate-, and high-mortality tertiles were assessed using the Kruskal-Wallis rank-sum test.

When the Kruskal-Wallis test indicated a statistically significant overall difference, pairwise comparisons were conducted using Dunn multiple comparison tests with Bonferroni adjustment to identify specific between-tertile differences. All statistical tests were two-sided with a significance threshold of p < 0.05.

### Ethical review

This study used publicly available, deidentified secondary data and was therefore exempt from institutional review board review.

## Results

County-level data availability varied by group and cancer type due to suppression for small counts. A total of 1,608 counties were included in the Non-Hispanic White breast cancer analysis, 1,434 in the Non-Hispanic White prostate cancer analysis, 344 in the Non-Hispanic Black breast cancer analysis, 303 in the Non-Hispanic Black prostate cancer analysis, 157 in the Hispanic breast cancer analysis, and 114 in the Hispanic prostate cancer analysis. Counties with missing or suppressed mortality data were excluded. All counties included in the mortality analyses had complete socio-behavioral determinant data.

The analytic sample represented a substantial proportion of the U.S. population for each racial and ethnic group studied, with coverage varying by cancer type and group (eTable 2). The analytic sample represented the majority of each population group: for breast cancer, 94% of Non-Hispanic White, 84% of Non-Hispanic Black, and 78% of Hispanic women; for prostate cancer, 91% of Non-Hispanic White, 78% of Non-Hispanic Black, and 70% of Hispanic men (eTable 2). Across cancer types and groups, the largest share of the represented population resided in moderate-mortality counties. Represented populations were smaller for Hispanic and Non-Hispanic Black strata than for Non-Hispanic White strata, reflecting suppression and underlying population distribution (eTable 2).

Counties were grouped into mortality tertiles within each race/ethnicity-by-cancer stratum (Table 1). Across race/ethnicity-by-cancer strata, Non-Hispanic Black populations had the highest population-weighted, age-adjusted mortality rates for both breast and prostate cancer across all tertiles, while Hispanic populations experienced the lowest mortality (Table 1).

**Table 1.** Average age-adjusted cancer mortality rate by mortality tertile.

| Cancer type | Group | Population-weighted, age-adjusted mortality rate by county mortality tertile (deaths per 100k population, 2018-2022) |  |  |
| --- | --- | --- | --- | --- |
|  |  | Low | Moderate | High |
| Breast | Non-Hispanic Black | 22.5 | 27.5 | 32.8 |
| Breast | Hispanic | 11.1 | 14.2 | 17.5 |
| Breast | Non-Hispanic White | 16.8 | 20.1 | 24.0 |
| Prostate | Non-Hispanic Black | 30.6 | 39.1 | 50.0 |
| Prostate | Hispanic | 12.9 | 16.9 | 20.6 |
| Prostate | Non-Hispanic White | 15.3 | 19.1 | 23.5 |

*Calculated by authors using data from the National Cancer Institute based on SEER and the American Community Survey (4,13)*

Across groups, domain profiles, not only composite scores, distinguished high- from low-mortality counties. This was especially evident for Hispanic populations, where lower average mortality masked pronounced socio-behavioral risk in high-mortality counties.

### Breast cancer socio-behavioral needs across mortality tertiles

Overall socio-behavioral risk increased monotonically across breast cancer mortality tertiles for all racial and ethnic groups (Figure 1A). Among Non-Hispanic Black women, mean socio-behavioral risk increased from 55 in low-mortality counties to 65 in moderate-mortality counties and 69 in high-mortality counties. Among Hispanic women, socio-behavioral risk increased from 51 to 67 and 74. Among Non-Hispanic White women, corresponding values increased from 34 to 43 and 55. For both Non-Hispanic Black and Hispanic women, the largest absolute increase in socio-behavioral risk occurred between low- and moderate-mortality counties, whereas for Non-Hispanic White women, increases were similar in magnitude across both transitions.

**Figure 1.**
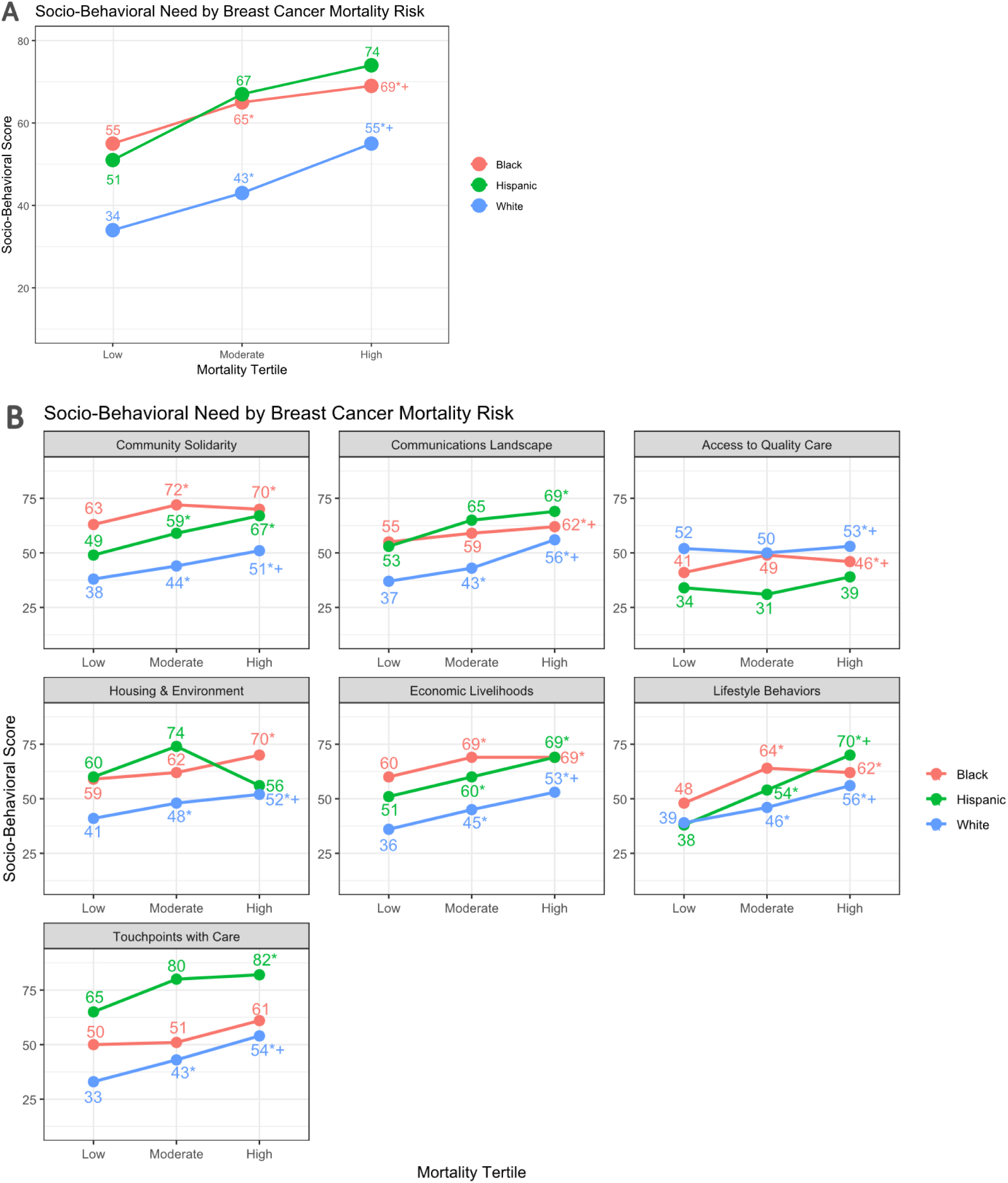
Socio-behavioral risk score by breast cancer mortality rate tertiles A) overall and B) by social needs category, per group (Non-Hispanic Black, Hispanic, Non-Hispanic White), 2018-2022. Symbols indicate statistically significant pairwise differences based on Dunn tests with Bonferroni adjustment (eTable 3). For moderate-mortality counties, * indicates a significant difference vs low-mortality counties. For high-mortality counties, * indicates a significant difference vs low-mortality counties, + indicates a significant difference vs moderate-mortality counties, and *+ indicates a significant difference vs both low- and moderate-mortality counties.

Domain-specific patterns varied by group (Figure 1B). Among Non-Hispanic Black women, the largest absolute differences between low- and high-mortality counties were observed for touchpoints with care, lifestyle behaviors, and community solidarity. Among Hispanic women, the largest absolute differences were observed for lifestyle behaviors, touchpoints with care, and economic livelihoods. Among Non-Hispanic White women, the domains with the largest absolute differences were touchpoints with care, economic livelihoods, and lifestyle behaviors.

Across all racial and ethnic groups, the domains showing the largest absolute gaps between high- and low-mortality counties were lifestyle behaviors, economic livelihoods, and community solidarity (Figure 1B). In high-mortality counties, the highest socio-behavioral risk for Non-Hispanic White women was concentrated in touchpoints with care, economic livelihoods, and lifestyle behaviors. For Non-Hispanic Black women, the highest needs were concentrated in touchpoints with care, lifestyle behaviors, and community solidarity. For Hispanic women, lifestyle behaviors, touchpoints with care, and economic livelihoods were the dominant contributors to elevated socio-behavioral need in high-mortality counties.

### Prostate cancer

Overall socio-behavioral risk generally increased across prostate cancer mortality tertiles for all racial and ethnic groups (Figure 2A). Among Non-Hispanic Black men, mean socio-behavioral risk increased from 54 in low-mortality counties to 65 in moderate-mortality counties and 66 in high-mortality counties. Among Hispanic men, socio-behavioral risk increased from 64 to 66 and 69 across the same tertiles. Among Non-Hispanic White men, corresponding values increased from 38 to 41 and 47. Gradients were less monotonic for prostate cancer than for breast cancer, particularly among Hispanic and Non-Hispanic White men.

**Figure 2.**
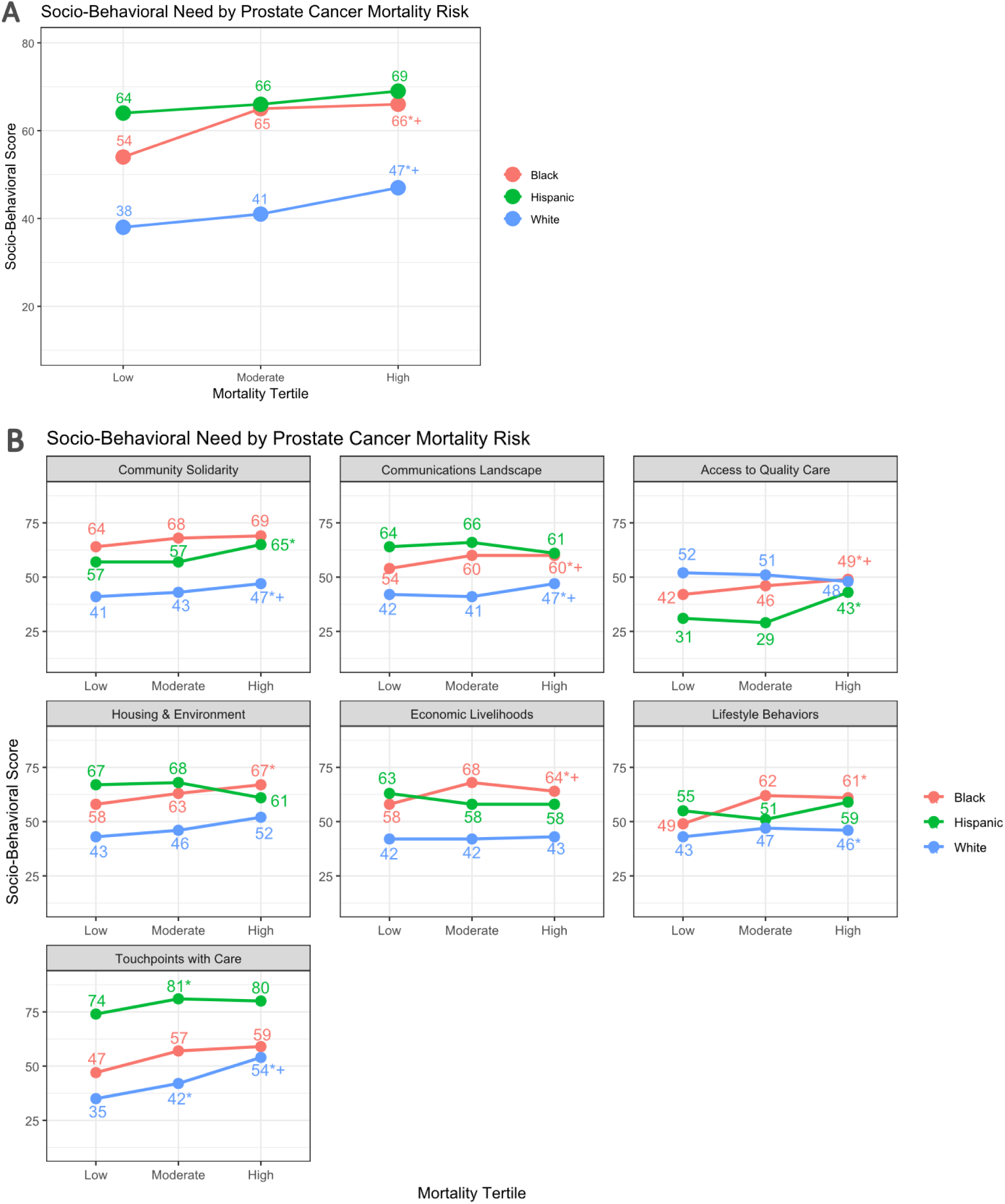
Socio-behavioral risk score by prostate cancer mortality rate tertiles A) overall and B) by category, per group (Non-Hispanic Black, Hispanic, Non-Hispanic White), 2018-2022. Note: Symbols indicate statistically significant pairwise differences based on Dunn tests with Bonferroni adjustment (eTable 3). For moderate-mortality counties, * indicates a significant difference vs low-mortality counties. For high-mortality counties, * indicates a significant difference vs low-mortality counties, + indicates a significant difference vs moderate-mortality counties, and *+ indicates a significant difference vs both low- and moderate-mortality counties.

Domain-specific analyses demonstrated heterogeneity in socio-behavioral patterns across mortality tertiles and racial and ethnic groups (Figure 2B). Among Non-Hispanic Black men, the largest absolute differences between low- and high-mortality counties were observed for economic livelihoods, lifestyle behaviors, and community solidarity. Among Hispanic men, the largest absolute differences were observed for touchpoints with care, economic livelihoods, and lifestyle behaviors. Among Non-Hispanic White men, the domains with the largest absolute differences were touchpoints with care, housing and environmental risks, and community solidarity.

Across all racial and ethnic groups, the domains showing the largest absolute gaps between high- and low-mortality counties were touchpoints with care and economic livelihoods (Figure 2B). In high-mortality counties, the highest socio-behavioral needs for Non-Hispanic Black men were concentrated in economic livelihoods, housing and environmental risks, and lifestyle behaviors. For Hispanic men, the highest needs were concentrated in touchpoints with care, housing and environmental risks, and economic livelihoods. For Non-Hispanic White men, the highest needs were concentrated in touchpoints with care, housing and environmental risks, and community solidarity.

## Discussion

In this national cross-sectional analysis of county-level socio-behavioral determinants of health and breast and prostate cancer mortality, we observed gradients in socio-behavioral need across mortality tertiles within Non-Hispanic Black, Non-Hispanic White, and Hispanic populations. Overall mortality patterns were consistent with prior literature, including higher mortality among Non-Hispanic Black populations and lower mortality among Hispanic and Non-Hispanic White populations (6), but this study moves beyond these averages by quantifying within-group heterogeneity in socio-behavioral vulnerability. By constructing race- and ethnicity-specific socio-behavioral indices and examining seven distinct domains, we show that communities with similar racial and ethnic compositions can have markedly different socio-behavioral risk and cancer mortality risk.

This work adds three main innovations to the existing evidence base. First, it focuses explicitly on within-group comparisons, rather than only contrasting racial and ethnic groups; it demonstrates that high-need communities exist within every group. Second, it uses race- and cancer-specific, domain-level socio-behavioral profiles, rather than a single deprivation score (7), to identify which types of barriers are most strongly associated with elevated mortality in different populations and geographies. Third, it applies this approach simultaneously to breast and prostate cancer, showing that socio-behavioral gradients are cancer-specific, with more monotonic patterns for breast cancer and more complex patterns for prostate cancer. Together, these innovations establish a practical analytic framework that links race- and cancer-specific mortality stratification with domain-level socio-behavioral risk to inform where targeted cancer prevention and control interventions may be prioritized.

Domain-level analyses showed that it is not only the overall level of socio-behavioral risk that matters, but also its composition. For breast cancer, the largest contrasts between high- and low-mortality counties were observed in lifestyle behaviors, economic livelihoods, community solidarity, and touchpoints with care. For prostate cancer, touchpoints with care and economic livelihoods were most prominent, while other domains showed weaker or nonmonotonic patterns. These results are consistent with prior evidence linking access to care and socioeconomic conditions to breast and prostate cancer outcomes (8), but the present study extends this literature by providing race- and cancer-specific domain profiles across mortality strata. Instead of a single deprivation gradient, high-mortality counties exhibit different socio-behavioral risk profiles across domains and populations, which is precisely the type of information needed to design interventions that are tailored to the local barrier profile rather than applying uniform strategies across settings.

Although Hispanic populations exhibited the lowest overall breast and prostate cancer mortality, they demonstrated among the steepest within-group socio-behavioral gradients across mortality tertiles. This pattern is consistent with the Hispanic paradox (14–16) and suggests that lower aggregate mortality does not imply uniformly lower vulnerability across Hispanic communities. Potential explanations include protective social structures, behavioral factors, selective migration, and data limitations such as misclassification or incomplete follow-up. Importantly, high-mortality Hispanic counties exhibited pronounced socio-behavioral risk related to lifestyle behaviors, economic livelihoods, and touchpoints with care, underscoring the need to avoid interpreting lower population-average mortality as lower priority for intervention.

Within-group heterogeneity was also evident for Non-Hispanic Black and Non-Hispanic White populations. For Non-Hispanic Black populations, socio-behavioral risk remained elevated across all mortality strata, suggesting a persistently high baseline of adverse determinants even in lower-mortality counties and underscoring the need for sustained investment alongside targeted action in the highest-mortality areas. Among Non-Hispanic Whites, socio-behavioral risk was lower on average but still varied meaningfully across mortality tertiles, indicating that high-risk counties with distinct domain-specific barrier profiles exist within majority-White geographies as well. Together, these findings indicate that within-group heterogeneity is not unique to Hispanic populations but is a consistent feature across racial and ethnic groups, reinforcing the need for population-specific, place-based approaches rather than assumptions based on group averages.

Observed mortality differences may reflect contributions from both cancer incidence and survival following diagnosis. Because this study used mortality rather than linked incidence and survival data, the relative contributions of cancer risk, stage at diagnosis, and post-diagnosis survival could not be separated. Socio-behavioral factors identified here may therefore influence multiple stages of the cancer continuum, including prevention, early detection, treatment access, and survivorship (5).

The findings have direct implications for the design of cancer control strategies. Many current investment and intervention decisions rely on broad socioeconomic indices or between-group disparities. Our results demonstrate that such approaches are likely to overlook high-need communities within racial and ethnic groups and may fail to identify the specific socio-behavioral barriers most strongly associated with elevated mortality. The differentiated domain patterns observed here support the use of geographically targeted, domain-specific strategies that align intervention design with the dominant needs of high-mortality communities.

Several limitations warrant consideration. This analysis is ecological and cross-sectional, and associations cannot be inferred at the individual level. However, socio-behavioral risk scores were constructed using race and ethnicity-specific population-weighted aggregation of census tract-level indicators, rather than overall county averages, allowing us to characterize within-county heterogeneity across populations. As with all county-level analyses, these estimates assume relative uniformity of socio-behavioral conditions within counties for each group and may mask finer-scale neighborhood variation. Race and ethnicity classification in cancer registry data may be incomplete in some settings (17). Socio-behavioral indicators were drawn from multiple sources and years, which may introduce temporal misalignment, though community-level socio-behavioral conditions generally change gradually over time. Suppression of mortality data for low-count counties reduced population coverage for some groups. Finally, the socio-behavioral index does not capture all structural or biological contributors to cancer mortality.

This study has the following strengths: national coverage, race- and ethnicity-specific mortality stratification, population-weighted estimation, and integration of multiple socio-behavioral domains. By quantifying both overall and domain-specific socio-behavioral gradients within racial and ethnic groups across mortality strata, this work provides a nuanced framework for understanding how structurally patterned vulnerability relates to cancer outcomes in the United States.

This socio-behavioral profiling approach provides an analytic framework for funders, health systems, and policy makers to guide cancer prevention and control investments. By jointly stratifying jurisdictions by race- and cancer-specific mortality and examining domain-level socio-behavioral risk profiles within high-mortality areas, the framework shifts decision-making from population averages to locally differentiated needs. This enables stakeholders to identify where mortality risk is greatest, but which specific socio-behavioral domains—such as touchpoints with care, economic livelihoods, or lifestyle behaviors—are most likely to constrain impact in a given population and place. Compared with approaches that rely solely on race, geography, or composite deprivation indices, this framework supports more precise alignment of intervention strategies with the dominant barriers faced by high-mortality communities, supporting more efficient and equitable cancer control investments.

## Conclusion

In this national county-level analysis, socio-behavioral determinants of health varied substantially across breast and prostate cancer mortality strata within Non-Hispanic Black, Non-Hispanic White, and Hispanic populations. Elevated mortality was consistently associated with higher and compositionally distinct socio-behavioral risk. By combining race- and cancer-specific mortality with local, domain-level socio-behavioral profiles, this study moves beyond traditional between-group comparisons and single deprivation scores to provide an implementation-relevant, data-driven approach for action. These findings highlight the importance of using accurate, community-specific socio-behavioral data to inform cancer prevention and control and offer a practical basis for designing more differentiated, equity-oriented intervention strategies that target the right domains in the right places.

## Data Availability

All behavioral data in the study are available to license following a request to the corresponding author.

## Supplement

### eMethods

#### Study sample

This study analyzed 3,141 U.S. counties, stratified into low-, moderate- and high-mortality tertiles for breast and prostate cancer among Non-Hispanic White, Non-Hispanic Black, and Hispanic populations. The total number of counties included in each racial/ethnic and cancer type subgroup varied due to differences in data availability and case counts. The number of counties analyzed for each group was:

- Non-Hispanic White Breast Cancer (W-BC): *n = 1,608 counties*
- Non-Hispanic White Prostate Cancer (W-PC): *n = 1,434 counties*
- Non-Hispanic Black Breast Cancer (AA-BC): *n = 344 counties*
- Non-Hispanic Black Prostate Cancer (AA-PC): *n = 303 counties*
- Hispanic Breast Cancer (H-BC): *n = 157 counties*
- Hispanic Prostate Cancer (H-PC): *n = 114 counties*

Counties with missing cancer mortality data were excluded from analysis. All counties included in the mortality analysis had complete socio-behavioral data.

#### Race and ethnicity definitions

We used the race and ethnicity definitions employed by the Census Bureau and established by the Office of Management and Budget (OMB) (1,2). Two racial groups are amongst the groups included in this study:

- White – A person having origins in any of the original peoples of Europe, the Middle East, or North Africa.
- Black or African American – A person having origins in any of the Black racial groups of Africa.

The OMB standards classify individuals in one of two ethnicity categories: “Hispanic or Latino” or “Not Hispanic or Latino.” OMB defines "Hispanic or Latino" as a person of Cuban, Mexican, Puerto Rican, South or Central American, or other Spanish culture or origin regardless of race. Throughout this article, we use the term “Hispanic or Latino” interchangeably with the term “Hispanic,” and refer to this concept as “ethnicity.” The OMB standards also state that people of Hispanic origin may be of any race.

In our analysis, we calculate the Hispanic or Latino population of any race as a category; and each of the race alone, non-Hispanic groups as individual categories:

- Hispanic: A person of Cuban, Mexican, Puerto Rican, South or Central American, or other Spanish culture or origin, regardless of race.
- White, non-Hispanic: A person having origins in any of the original peoples of Europe, the Middle East, or North Africa, and of non-Hispanic ethnicity.
- Black or African American, non-Hispanic: A person having origins in any of the Black racial groups of Africa, and non-Hispanic ethnicity.

Because Hispanic or Latino origin is classified as an ethnicity rather than a race, the Hispanic population in this study includes individuals of any racial background and therefore represents a heterogeneous group with respect to racial identity.

#### Index Calculation

The index of socio-behavioral risk to adverse health outcomes is calculated by aggregating socio-behavioral indicators that reflect community well-being, access to resources, and health system engagement. These indicators are organized into seven key themes: community solidarity, education, health literacy, and connectivity, quality care, housing and environmental risks, economic livelihoods, lifestyle behaviors, and touchpoints with care.

1. Community Solidarity: This theme measures the strength of social support systems within a community. Indicators include adult incarceration rates, limited access to childcare facilities, low childcare worker density, SNAP recipients (poverty-adjusted), shortages of SNAP-authorized retailers, disconnected youth (youth not in school or employed), and residential segregation metrics like the dissimilarity index and the diversity index. Single-parent households are also included, as they often face higher socio-behavioral risks.
2. Education, Health Literacy, and Connectivity: Educational attainment and access to health information shape a community’s ability to engage with health services. Indicators such as incomplete high school education, low literacy and numeracy, no household internet or smartphone, and populations with limited English proficiency capture barriers to information access. Health literacy measures include whether adults understand clinical instructions and access medical records online.
3. Quality Care: This theme evaluates access to healthcare services. Indicators include the availability of hospitals, pharmacies, and urgent care centers, hospital bed density, PQI composite scores (preventable hospital admissions), median emergency department wait times, and the provision of appropriate sepsis care. These measures reflect the quality and accessibility of care within a community.
4. Housing and Environmental Risks: Secure housing and environmental safety are critical for health. Indicators include household crowding, high concentrations of group quarters, insufficient housing (mobile homes, multi-unit structures with incomplete kitchens or plumbing), high housing costs (households spending over 50% of income on housing), and the proportion of renters. Environmental risks include air pollution, economic losses from natural hazards, long commutes, solitary commuting, vehicle ownership, and distance to transit stops.
5. Economic Livelihoods: Economic stability is a major determinant of health. Indicators such as low median household income, poverty, income inequality, unemployment, and the number of available jobs per household provide insight into economic vulnerability. Additional indicators include the proportion of low-wage jobs, job-to-skill mismatches, lack of health insurance, and medical debt in collections.
6. Lifestyle Behaviors: Health outcomes are influenced by individual lifestyle choices. This theme includes tobacco and alcohol use, low physical activity levels, limited access to parks and recreational facilities, limited access to supermarkets, and high fast-food consumption—factors that contribute to poor health outcomes.
7. Touchpoints with care: Regular engagement with healthcare services is essential for preventing adverse health outcomes. Indicators include lack of a dedicated provider, missed regular check-ups, no annual dental visits, and gaps in preventive care screening for conditions like high blood pressure, cholesterol, colorectal cancer, and cervical cancer. This theme also tracks core preventive services among older adults, such as flu and pneumococcal vaccinations.

Each indicator is assigned a percentile ranking relative to all other counties in the U.S. These percentile rankings are then aggregated to produce 7 thematic and a single composite index of socio-behavioral vulnerability. Higher index scores signify communities more at risk of adverse health outcomes due to socio-behavioral and environmental factors. This index is a critical tool for identifying vulnerable populations and targeting public health interventions

#### Calculation of socio-behavioral score by cancer mortality tertile

We computed county-level socio-behavioral determinants of health (SBDOH) scores separately for each race/ethnicity-by-sex group by aggregating census tract–level SBDOH scores using tract population estimates for that same group as weights. Specifically, for each county and group, we calculated a population-weighted mean of tract SBDOH scores, restricted to tracts within the county. Because scores are constructed within group (using group-specific population weights), they are intended to characterize *within-group* geographic variation and should not be used to directly compare absolute SBDOH levels across groups.

Let:

- t index census tracts in county c
- g index race/ethnicity-by-sex group (e.g., Black males)
- S_t_ be the tract-level SBDOH Need Index (or theme) score for tract t
- Pop_t,g_ be the estimated adult population for group g in tract t Then the county-level SBDOH score for county c and group g is:

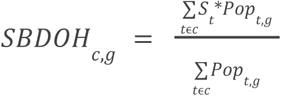

**eTable 1.** Social and Behavioral Determinants of Health (SBDOH) variables included in the analysis, by category.

| Category | Measures |
| --- | --- |
| Community Solidarity | Adult incarceration<br>Limited access to childcare facilities<br>Low childcare facility worker density<br>SNAP recipients (poverty-adjusted)<br>Shortage of SNAP-authorized retailers<br>Disconnected youth (out of school and unemployed)<br>Residential segregation: Dissimilarity index<br>Diversity index (living among communities of same race)<br>Single-parent household |
| Education, Health Literacy, and Connectivity | Incomplete high school education<br>Low literacy<br>Low numeracy<br>No household internet<br>No household smartphone or computer<br>Population with limited or no English<br>Health literacy: Adults whose provider asks for understanding of clinical instructions<br>Health literacy: Adults who access their medical records online |
| Quality Care | Limited access to hospitals<br>Limited access to pharmacies<br>Limited access to urgent care centers<br>Low availability of hospital beds<br>PQI Composite (admission rate for preventable conditions)<br>Average (median) emergency department visit time<br>Appropriate sepsis care |
| Housing and Environmental Risks | Household crowding<br>High concentration of group quarters<br>Insufficient housing (mobile homes, multi-unit, incomplete kitchen or plumbing)<br>Households spending over 50% of income on housing<br>Population who rent their housing (instead of own)<br>Air pollution<br>High estimated economic loss due to natural hazards<br>Long commute (> 30 minutes)<br>Workers commuting alone<br>No vehicle ownership<br>Distance to nearest transit stop |
| Economic Livelihoods | Low median household income<br>Poverty<br>Income inequality<br>Unemployment<br>Few available jobs per household<br>High share of low or living wage jobs<br>Job-to-skill imbalance<br>Lack of health insurance coverage<br>Medical debt in collections |
| Lifestyle Behaviors | Tobacco use<br>Alcohol use<br>Low physical activity<br>Limited access to parks and recreational centers<br>Limited access to supermarkets<br>High fast-food consumption |
| Touchpoints with care | Lack of a dedicated provider or usual source of care<br>Failure to complete regular check-ups<br>No annual dental care visit<br>Uncontrolled blood pressure<br>Lack of preventive care screening for cholesterol among adults 18+<br>Lack of preventive care screening for colorectal cancer among adults 50-75<br>Lack of preventive care screening for cervical cancer among women 21-65<br>Lack of core preventive services (flu shot, ppsv shot, colorectal cancer screening) among women age 65+<br>Lack of core preventive services (flu shot, ppsv shot, colorectal cancer screening) among men age 65+ |

**eTable 2.** Population and percentage of the population by cancer incidence rate tertile.

| Cancer type | Group | Proportion of population in the U.S. represented in study (%) | Population in millions (proportion of US population) by age-adjusted cancer mortality tertile (2018-2022) |  |  |
| --- | --- | --- | --- | --- | --- |
|  |  |  | Low | Moderate | High |
| Breast | Non-Hispanic Black | 84% | 4.5 (28%) | 5.7 (35%) | 3.3 (21%) |
| Breast | Hispanic | 78% | 4.5 (22%) | 7.5 (37%) | 4.0 (20%) |
| Breast | Non-Hispanic White | 94% | 27 (35%) | 30 (40%) | 14 (19%) |
| Prostate | Non-Hispanic Black | 78% | 3.9 (28%) | 5.2 (36%) | 2.0 (14%) |
| Prostate | Hispanic | 70% | 4.7 (23%) | 6.8 (33%) | 3.1 (15%) |
| Prostate | Non-Hispanic White | 91% | 30 (41%) | 22 (30%) | 15 (20%) |

**eTable 3.** Association between socio-behavioral needs and cancer mortality by race and ethnicity.

| Theme | Cancer type | Race and Ethnicity | Normality (p-value) | Kruskal (p-value)<sup>a</sup> | Dunn High vs Low (p-value) | Dunn High vs Moderate (p-value) | Dunn Moderate vs Low (p-value) |
| --- | --- | --- | --- | --- | --- | --- | --- |
| Index | Breast | White | 9.54E-11 | NA | 1.27E-45 | 3.14E-45 | 1.38E-19 |
| Theme1 | Breast | White | 0.004559 | NA | 1.59E-19 | 4.70E-20 | 7.20E-07 |
| Theme2 | Breast | White | 1.89E-16 | NA | 1.87E-39 | 4.88E-38 | 5.26E-20 |
| Theme3 | Breast | White | 5.67E-11 | NA | 1.98E-06 | 2.07E-06 | 0.000929 |
| Theme4 | Breast | White | 1.52E-13 | NA | 5.64E-09 | 2.17E-09 | 0.008964 |
| Theme5 | Breast | White | 1.32E-15 | NA | 2.84E-33 | 1.55E-33 | 4.77E-13 |
| Theme6 | Breast | White | 3.48E-13 | NA | 3.81E-25 | 2.60E-25 | 1.33E-10 |
| Theme7 | Breast | White | 7.63E-09 | NA | 3.86E-27 | 1.51E-27 | 2.60E-10 |
| Index | Breast | Black | 1.12E-07 | NA | 3.61E-10 | 1.40E-10 | 0.009906 |
| Theme1 | Breast | Black | 7.91E-10 | NA | 6.76E-06 | 1.43E-05 | 1 |
| Theme2 | Breast | Black | 5.06E-05 | NA | 2.74E-08 | 1.41E-08 | 0.001101 |
| Theme3 | Breast | Black | 0.000233 | NA | 0.000323 | 0.000221 | 0.036159 |
| Theme4 | Breast | Black | 4.34E-07 | NA | 0.015501 | 0.012065 | 0.65237 |
| Theme5 | Breast | Black | 1.23E-10 | NA | 8.32E-08 | 8.48E-08 | 0.307061 |
| Theme6 | Breast | Black | 2.82E-10 | NA | 1.45E-06 | 1.22E-06 | 0.337584 |
| Theme7 | Breast | Black | 0.000851 | NA | 0.232517 | 0.288532 | 1 |
| Index | Breast | Hispanic | 0.284056 | 1.94E-07 | NA | NA | NA |
| Theme1 | Breast | Hispanic | 3.81E-05 | NA | 1.56E-05 | 8.60E-06 | 0.102881 |
| Theme2 | Breast | Hispanic | 0.002794 | NA | 0.000327 | 0.000185 | 0.081051 |
| Theme3 | Breast | Hispanic | 0.012052 | NA | 0.13226 | 0.174391 | 1 |
| Theme4 | Breast | Hispanic | 0.000332 | NA | 0.824752 | 1 | 1 |
| Theme5 | Breast | Hispanic | 0.023668 | NA | 4.68E-06 | 2.44E-06 | 0.070618 |
| Theme6 | Breast | Hispanic | 0.015864 | NA | 1.77E-08 | 7.55E-09 | 0.012882 |
| Theme7 | Breast | Hispanic | 3.44E-05 | NA | 0.000213 | 0.000121 | 0.126596 |
| Index | Prostate | White | 1.93E-08 | NA | 1.69E-09 | 4.92E-09 | 5.99E-06 |
| Theme1 | Prostate | White | 0.002776 | NA | 4.44E-05 | 0.000262 | 0.000385 |
| Theme2 | Prostate | White | 4.32E-13 | NA | 2.68E-08 | 1.94E-07 | 5.82E-06 |
| Theme3 | Prostate | White | 4.39E-09 | NA | 0.059998 | 1 | 0.080344 |
| Theme4 | Prostate | White | 4.53E-12 | NA | 0.569892 | 1 | 1 |
| Theme5 | Prostate | White | 1.14E-13 | NA | 0.434826 | 1 | 0.66166 |
| Theme6 | Prostate | White | 4.03E-11 | NA | 0.03171 | 0.025843 | 0.598709 |
| Theme7 | Prostate | White | 3.05E-09 | NA | 3.02E-29 | 1.40E-29 | 3.48E-11 |
| Index | Prostate | Black | 1.79E-05 | NA | 1.35E-05 | 6.68E-06 | 0.034485 |
| Theme1 | Prostate | Black | 6.46E-09 | NA | 0.051611 | 0.057207 | 0.244592 |
| Theme2 | Prostate | Black | 0.001858 | NA | 0.000208 | 0.000243 | 0.007986 |
| Theme3 | Prostate | Black | 0.000324 | NA | 0.000797 | 0.000633 | 0.03846 |
| Theme4 | Prostate | Black | 7.58E-05 | NA | 0.015995 | 0.013244 | 0.853182 |
| Theme5 | Prostate | Black | 1.36E-09 | NA | 0.000118 | 6.87E-05 | 0.042171 |
| Theme6 | Prostate | Black | 3.65E-09 | NA | 0.00079 | 0.000473 | 0.143251 |
| Theme7 | Prostate | Black | 0.00127 | NA | 0.48218 | 1 | 1 |
| Index | Prostate | Hispanic | 0.238478 | 0.035794 | NA | NA | NA |
| Theme1 | Prostate | Hispanic | 0.000142 | NA | 0.014958 | 0.01914 | 0.076055 |
| Theme2 | Prostate | Hispanic | 0.031572 | NA | 0.568725 | 0.867729 | 1 |
| Theme3 | Prostate | Hispanic | 0.002313 | NA | 0.002792 | 0.002152 | 0.645625 |
| Theme4 | Prostate | Hispanic | 0.001933 | NA | 0.145644 | 0.190801 | 0.402252 |
| Theme5 | Prostate | Hispanic | 0.059377 | 0.77463 | NA | NA | NA |
| Theme6 | Prostate | Hispanic | 0.044382 | NA | 0.065594 | 0.06731 | 1 |
| Theme7 | Prostate | Hispanic | 4.23E-05 | NA | 0.027034 | 0.080921 | 1 |

*Kruskal-Wallis and Dunn's ad-hoc p-values by socio-behavioral need category, cancer type, and race/ethnicity. <sup>a</sup>“NA” indicates that a post-hoc Dunn test was not performed because the overall Kruskal-Wallis test was not statistically significant; in such cases, pairwise comparisons are not applicable. A significant p-value in the “Normality” column refers to rejection of the null hypothesis of the normality test (i.e., evidence against a normal distribution), which justifies our choice of nonparametric methods (Kruskal-Wallis and Dunn).*

## Notes

### Competing Interest Statement

Several authors are employees of Surgo Health, a public benefit corporation that holds the right to the socio-behavioral index used in this manuscript.

### Funding Statement

This study was supported by Surgo Health P.B.C. and Pfizer, Inc.. No external grant numbers were assigned for this work. Support from Surgo Health included funding for data collection and contribution to manuscript preparation. Pfizer provided additional financial support.

### Author Declarations

Age-adjusted breast and prostate cancer mortality rates stratified by race and ethnicity for the period 2018-2022 were obtained from the National Cancer Institute.

### Summary of Updates

Corrected the author order which contained a mistake.

